# Molecular typing and characterisation of antimicrobial resistance determinants of *Neisseria gonorrhoeae* among symptomatic individuals in the Eastern Cape, South Africa

**DOI:** 10.64898/2026.11.06.26367404

**Authors:** Mildred Mandisa Mdingi, Brian Kullin, Hyunsul Jung, Nireshni Mitchev, Remco P.H Peters

## Abstract

**Background:** Antimicrobial resistance in *Neisseria gonorrhoeae* threatens gonorrhoea treatment, while surveillance is constrained by limited culture capacity. We evaluated multi-antigen sequence typing (NG-MAST) and targeted real-time PCR for molecular epidemiology and detection of antimicrobial resistance-associated determinants directly from clinical specimens.

**Methods:** We analysed 235 *N. gonorrhoeae*-positive stored DNA extracts from symptomatic individuals in South Africa. NG-MAST sequence types were obtained directly or from whole-genome sequences (WGS) of paired cultured isolates. Real-time PCR targeted fluoroquinolone resistance-associated mutations (RAMs) in *parC* and *gyrA* and macrolide RAMs in the 23S rRNA gene. Results were compared with WGS and with phenotypic susceptibility results from paired isolates.

**Results:** NG-MAST sequence types were obtained for 200/235 (85.1%) specimens and were diverse, with ST21253 predominating (n=26). At least one *gyrA* or *parC* mutation was detected by real-time PCR in 110/193 (57.0%) specimens. Compared with WGS, positive agreement was 81.2% for *parC* and 97.0% for *gyrA*, and negative agreement 100% and 95.8%, respectively. Among 65 ciprofloxacin-resistant isolates, RAMs were detected in 60 (92.3%) by real-time PCR and 61 (93.8%) by WGS; four had no RAM detected by either method. The 23S rRNA C2611T mutation was detected by both methods in one isolate (azithromycin MIC, 8 mg/L); no A2059G mutations were detected.

**Conclusions:** Gonococcal sequence types were diverse, fluoroquinolone RAMs were common, and the assessed macrolide RAMs were rare. Real-time PCR applied directly to specimen DNA agreed well with WGS, but the targeted markers did not fully predict ciprofloxacin susceptibility, supporting complementary molecular and phenotypic approaches.

## INTRODUCTION

The management and control of gonorrhoea have become increasingly challenging because of the emergence and spread of antimicrobial-resistance (AMR) in *Neisseria gonorrhoeae*, which threaten the effectiveness of available treatment options.(1) Historically, *N. gonorrhoeae* has developed resistance to multiple antimicrobial classes, including sulphonamides, penicillins, tetracyclines, and fluoroquinolones.(1) More recently, reduced susceptibility and resistance to extended-spectrum cephalosporins and azithromycin have raised concerns about the sustainability of current treatment strategies.(2) Consequently, World Health Organization (WHO) has identified antimicrobial-resistant *N. gonorrhoeae* as a priority pathogen that requires strengthened antimicrobial resistance (AMR) surveillance and innovative approaches to detect and monitor emerging resistance.(3)

Phenotypic antimicrobial susceptibility testing (AST) remains central to gonococcal AMR surveillance, but requires successful culture of viable isolates, specialised laboratory infrastructure and technical expertise. (4, 5) These requirements present particular challenges in low- and middle-income countries (LMICs), including settings where syndromic management is the standard of care and gonococcal culture capacity is limited.(5–7)

Molecular methods that detect resistance-associated mutations (RAMs) directly from *N. gonorrhoeae*-positive clinical specimens provide a complementary approach to culture-based surveillance, particularly in settings where culture facilities are limited or not routinely available.(5, 6) Molecular characterisation can also provide information on gonococcal genetic diversity and the circulation and spread of AMR-associated lineages.(5, 8, 9) *N. gonorrhoeae* multi-antigen sequence typing (NG-MAST), based on sequence variation in internal fragments of the highly polymorphic *porB* and *tbpB* genes, has been widely used to characterize gonococcal strain diversity and circulating genotypes.(5, 9)

Fluoroquinolone resistance in *N. gonorrhoeae* is primarily associated with mutations within the fluoroquinolone resistance-determining regions of the *gyrA* and *parC* genes. Mutations at codons 91 and 95 of *gyrA* are strongly associated with ciprofloxacin resistance, and together with *parC* mutations can contribute to higher minimum inhibitory concentration (MIC) levels. Detection of fluoroquinolone-RAMs may therefore support molecular surveillance and potentially inform clinicians to implement resistance-guided ciprofloxacin treatment.(1, 10, 11) Azithromycin resistance in *N. gonorrhoeae* is associated with several resistance mechanisms, including mutations in the 23S rRNA gene that affect the macrolide binding site and increased drug efflux mediated by the *MtrCDE* pump. The A2059G and C2611T mutations have been associated with reduced azithromycin susceptibility and, depending on the specific mutation and number of affected 23S rRNA alleles, increased azithromycin MICs.(1, 12, 13)

Despite the public health importance of gonococcal AMR, molecular data describing circulating *N. gonorrhoeae* strains and AMR determinants in South Africa remain limited due to the limited culture capacity. This study aimed to characterise the molecular epidemiology and fluoroquinolone and macrolide AMR determinants of *N. gonorrhoeae* among symptomatic individuals in South Africa. Specifically, we determined (i) the distribution of the NG-MAST sequence types, (ii) the frequency of fluoroquinolone and macrolide RAMs by multiplex real-time PCR applied directly to stored specimen DNA, and (iii) agreement between targeted real-time PCR and whole genome sequencing of paired cultured isolates, and (iv) the relationship between detected mutations and phenotypic ciprofloxacin and azithromycin susceptibility.

## MATERIALS AND METHODS

### Clinical sample collection

This retrospective analysis used stored *N. gonorrhoeae*-positive clinical specimens, collected during a previously conducted cross-sectional diagnostic evaluation study.(14) The parent study evaluated a novel lateral flow assay for the detection of *N. gonorrhoeae* among symptomatic men and women attending five public healthcare clinics in Buffalo City Municipality, Eastern Cape, South Africa, between March and September 2022.(14) In the parent study, urine specimens were collected from men and nurse-collected vaginal swabs from women. Specimens were divided for lateral flow assay testing, Xpert CT/NG testing and storage. Testing using the Xpert CT/NG assay (Cepheid, Sunnyvale, CA, USA) was performed at the point of care, and specimens positive for *N. gonorrhoeae* and/or *Chlamydia trachomatis* were archived at 4°C at the Department of Medical Microbiology, University of Pretoria until DNA extraction.(14) The DNA was subsequently extracted using the High Pure PCR Template Preparation Kit (Roche Diagnostics GmbH, Germany) and was stored at - 20°C. For the present analysis, stored DNA extracts from 235 *N. gonorrhoeae*-positive specimens were included.

The study was approved by the University of Pretoria, Faculty of Health Sciences (UP HREC ref: 42/2024). All laboratory analyses were conducted at the Department of Medical Microbiology, University of Pretoria.

### *Neisseria gonorrhoeae* multiantigen sequence typing (NG-MAST)

Extracted DNA was subjected to PCR amplification of the *porB* and *tbpB* genes using previously described primers(15) and Celtaq HS DNA Polymerase (Celtic Molecular Diagnostics, South Africa). PCR amplification was performed using a SimpliAmp^™^

Thermal Cycler (ThermoFisher Scientific Inc., USA) under the following conditions: (i) pre-incubation: 95°C for 5 min; (ii) amplification: 40 cycles of 95°C for 30 s, 58°C (*porB*) or 69°C (*tbpB*) for 1 min, and 68°C for 2 min 30 s; and (iii) final extension: 68°C for 10 min. Expected amplicon sizes were 1,233 bp for *porB* and 1854 bp to 1875 bp for *tbpB*. Amplicons were confirmed by 1% agarose gel electrophoresis, purified using ExoSAP-IT™ (ThermoFisher Scientific Inc., USA), and subjected to bidirectional Sanger sequencing at Inqaba Biotechnical Industries (Pty) Ltd, South Africa.

### Molecular detection of resistance-associated mutations by multiplex real-time PCR

Stored DNA extracts were analysed using previously described multiplex real-time PCR assays to detect mutations associated with ciprofloxacin and macrolide resistance (Supplementary Table 1).(10) The assay targeted fluoroquinolone resistance-associated mutations in *parC* (any of D86N, S87R, S87N, S87I, S88P) and *gyrA* (any of S91F, S91P, S91Y), and macrolide RAMs at positions A2059 and C2611 in the 23S rRNA gene (*E. coli* numbering).

The real-time PCR reaction mixture was prepared in a final volume of 20 μL using TaqMan^®^ Multiplex Master Mix (Thermo Fisher Scientific, Inc., USA), TaqMan^®^ minor groove binding (MGB) probe or TaqMan™ probes with a QSY^®^ quencher (Thermo Fisher Scientific Inc., USA), primers (Inqaba Biotechnical Industries (Pty) Ltd, South Africa) and nuclease-free water according to the manufacturer’s instructions. Each PCR reaction consisted of 19 μL reaction mixture and 1 μL of DNA extract. *N. gonorrhoeae* ATCC 49226 culture was included as a positive control, and a negative control (nuclease-free water instead of DNA) was included in each run. Samples that failed the first run were retested using the same DNA extracts.

Amplification was performed using a QuantStudio™ 5 Real-Time PCR System (Thermo Fisher Scientific Inc., USA). The cycling conditions comprised an initial incubation at 60°C for 30 seconds and 95°C for 5 minutes, followed by 45 cycles at 95°C for 15 seconds and 60°C for 1 minute. An amplification with a typical sigmoidal curve and a cycle quantification (Cq) value below 40 were considered to be successfully amplified.

### Whole genome sequencing

Paired-end WGS data from cultured *N. gonorrhoeae* isolates corresponding to clinical specimens included in the present study were available from a previously published study.(16) In brief, WGS of *N. gonorrhoeae* isolates was performed using the paired-end (150 bp x 2) sequencing (NextSeq Illumina, USA), followed by *de novo* genome assembly using the Jekesa pipeline (https://github.com/stanikae/jekesa). Resistance-associated determinants in *gyrA*, *parC* and the 23S rRNA gene were identified from the assembled genomes using Pathogenwatch.(17)

### Antimicrobial susceptibility testing

Ciprofloxacin and azithromycin susceptibility testing results from cultured *N. gonorrhoeae* isolates corresponding to clinical specimens were available from the previously published study.(16) In brief, *N. gonorrhoeae* culture was performed on directly inoculated New York City agar with same-day CO₂-supported transport and incubation at 37°C in a CO₂-rich environment; presumptive colonies were identified by Gram staining, oxidase testing, and the API-NH strips. Antimicrobial susceptibility testing was performed using Etests (bioMérieux) according to the manufacturer’s instructions. MICs were interpreted according to EUCAST v12.0 (2022), with isolates classified as susceptible at ≤0.032 mg/L and resistant at >0.064 mg/L; isolates with an MIC of 0.064 mg/L were classified separately for descriptive purposes. The epidemiological cut-off (>1mg/L) was used for azithromycin resistance.

### Bioinformatics and statistical analysis

NG-MAST v2.0(18) was used to genotype all samples for which *porB* and *tbpB* nucleotide sequences were both available. Nucleotide sequences obtained from Sanger sequencing were analysed and assembled using the Staden Package version 2.0. For WGS data, *porB* and *tbpB* sequences were extracted using a custom python script. All sequences were then matched against the *N. gonorrhoeae* pubMLST database(19) https://pubmlst.org/bigsdb?db=pubmlst_neisseria_seqdef&page=profiles) to obtain the NG-MAST sequence types (STs). Allelic profiles that did not match any entries in the NG-MAST v2.0 pubMLST database were considered novel; these were submitted to the pubMLST database, where ST numbers were assigned. Following the ST assignment, alleles for each sample were aligned using mafft v7.525 (20) and concatenated in frame. Maximum likelihood phylogenies were generated for these alignments using IQTREE v3.1.4 (21) with 1000 bootstrap replicates and best fit gene-partitioned models predicted by the program. The resulting trees were viewed and edited using the iToL v7.6 tree viewer.

Descriptive statistics were used to summarize participant characteristics, NG-MAST sequence types, resistance-associated mutations and antimicrobial susceptibility results. Categorical variables were reported as frequencies and percentages, and continuous variables as medians and ranges. Proportions were calculated using the number of specimens or isolates with an available result for the respective analysis as the denominator.

Agreement between real-time PCR and WGS for detection of each targeted mutation was assessed using WGS as the reference method. Positive and negative agreement were calculated with exact binomial 95% confidence intervals. Positive agreement was defined as the proportion of samples identified to harbor any one of the targeted single nucleotide polymorphism (SNP) mutations at each gene position by real-time PCR to samples identified to have any corresponding SNP mutations by WGS, and negative agreement as the proportion of samples that were wild type at any one of the targeted gene position by real-time PCR to samples that were wild type at each targeted gene position by WGS. The relationship between detection of targeted RAMs and phenotypic antimicrobial susceptibility was assessed descriptively among isolates with paired molecular and phenotypic results.

## RESULTS

### Overview of sample population

A total of 235 *N. gonorrhoeae*-positive DNA extracts were included in the analysis. Of these, 167 (71.1%) were derived from urine specimens from men and 68 (28.9%) from vaginal swabs from women. The median participant age was 24 years (range 18 - 59 years). Overall, 27/235 (11.5%) participants were living with HIV, 170/235 (72.3%) tested HIV negative and 38/235 (16.1%) had unknown HIV status. Fourteen participants (6.0%) reported treatment for STI-associated symptoms in the previous month and 47 (20.0%) in the previous 12 months.

### Strain diversity of *N. gonorrhoeae* as determined by NG-MAST

NG-MAST sequence types (STs) were derived from WGS data for 104 (44.3%) *N. gonorrhoeae* isolates. Among isolates without an NG-MAST type derived from WGS data, NG-MAST typing was successful for 96/131 (73.3%) DNA extracts. Of the samples that failed to amplify the NG-MAST genes, 6/131 (4.6%) samples successfully amplified only one of two genes, while none was successfully amplified in 29/131 (22.1%).

A diverse range of NG-MAST STs was observed across the study population (Supplementary Table 2). ST21253 was the most frequently identified (n=26), followed by ST19216 and ST21254 (n=6 each), ST21062 (n=5), and ST2670, ST21047, and ST21195 (n=4 each). ST21253 was identified at all five participating facilities and among both men (n=21) and women (n=5). There was no apparent phylogenetic clustering by study site or gender (Figure 1).

**Figure 1.**
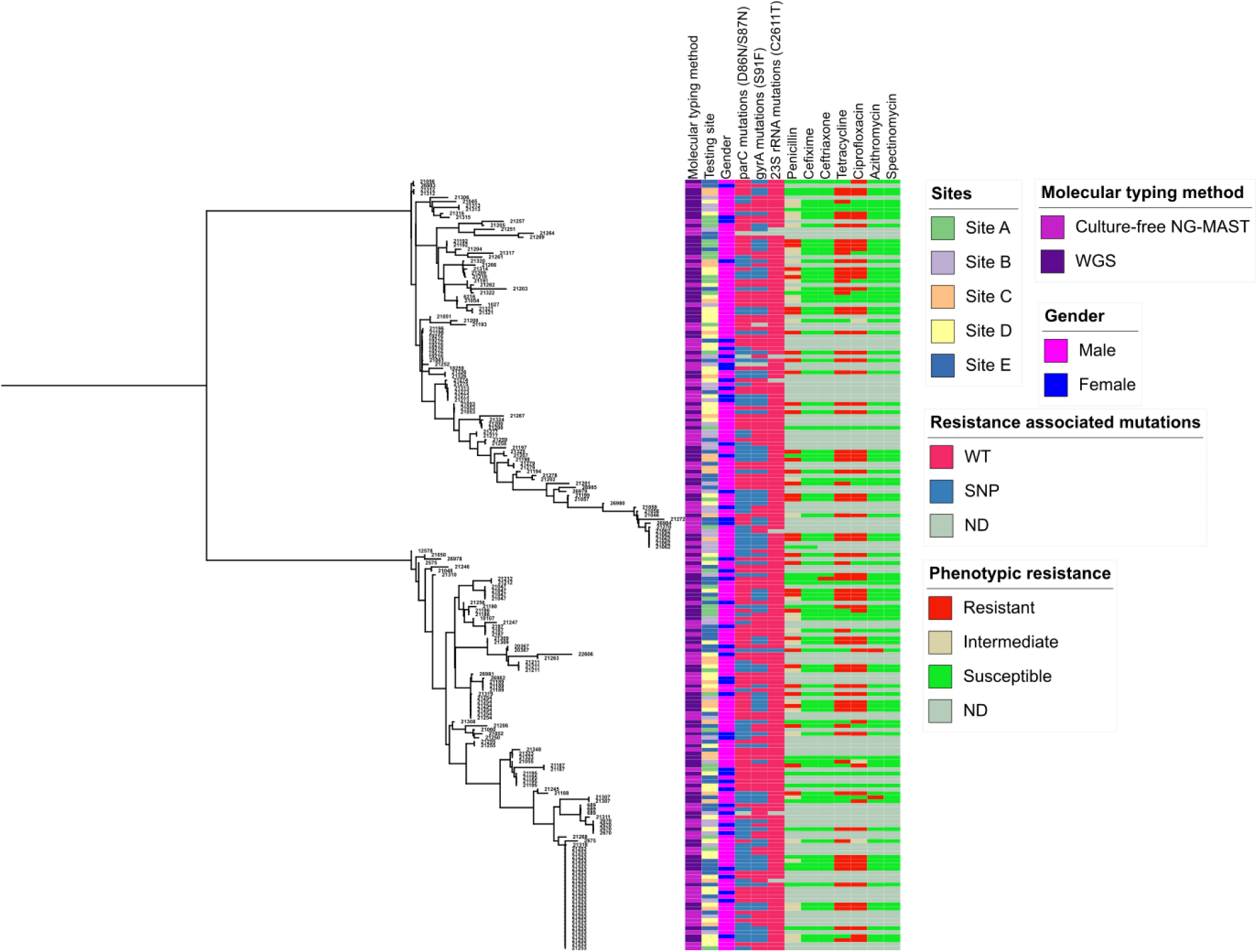
Phylogenetic relationships and antimicrobial resistance characteristics of *Neisseria gonorrhoeae* isolates from symptomatic individuals in the Eastern Cape, South Africa. **Note**. The phylogenetic tree was constructed using concatenated *porB* and *tbpB* sequences. Study site, participant sex, molecular typing method, NG-MAST sequence type (ST), and resistance-associated mutations are indicated alongside the tree. Resistance-associated mutations include *parC* D86N and S87N, *gyrA* S91F, and 23S rRNA C2611T. Phenotypic susceptibility is indicated as susceptible (S), intermediate (I), or resistant (R) for penicillin, cefixime, ceftriaxone, tetracycline, ciprofloxacin, azithromycin, and spectinomycin. WT, wild type; SNP, resistance-associated mutation detected; ND, not determined; WGS, whole-genome sequencing; NG-MAST, *N. gonorrhoeae* multi-antigen sequence typing.

### Distribution of resistance-associated mutations determined by real-time PCR and whole genome sequencing

Resistance-associated mutations in the *gyrA*, *parC* and 23S rRNA genes were successfully assessed by multiplex real-time PCR in 193/235 (82.1%) samples; amplification was unsuccessful in 42 (17.9%) samples. The frequency of targeted RAMs is summarised in (Table 1). At least one targeted *parC* or *gyrA* mutation was detected in 110/193 (57.0%) samples. Mutations in both genes were detected in 46 (23.8%) samples, while 32 (16.6%) had *parC* mutations only and 32 (16.6%) *gyrA* mutations only. One had an undetermined *parC* PCR result. The 23S rRNA C2611T mutation was detected in one sample (1/193, 0.5%); and no A2059G mutations were detected.

**Table 1.** Frequency of quinolone and macrolide resistance-associated mutations in *Neisseria gonorrhoeae* samples (n=193) as determined by multiplex real-time PCR assay.

| **Gene** | **Mutations detected at targeted site** | **Wild type at the targeted position** | **Undetermined** |
|---|---|---|---|
| *gyrA* S91 | 78* (40.4%) | 115 (59.6%) | 0 |
| *parC* D86, S87, S88 | 78* (40.4%) | 114 (59.1%) | 1 (0.5%) |
| *gyrA* and *parC* | 46 (23.8%) | 147 (76.2%) | 0 |
| 23S – A2059G | 0 | 191 (98.9%) | 2 (1.1%) |
| 23S – C2611T | 1 (0.5%) | 190 (98.4%) | 2 (1.1%) |

**Note.** Proportions are row proportions.
\*Forty-six samples had mutations in both *parC* and *gyrA*; 32 had a *parC* mutation only, 32 had a *gyrA* mutation only.

The WGS results were available from previous studies for 90/193 (46.6%) samples that were subjected to real-time PCR. For *parC*, real-time PCR detected mutations in 39/48 (81.2%) samples identified by WGS as non-wild-type and concordantly classified all 42/42 (100%) WGS wild-type samples (Table 2). By mutation, real-time PCR detected 1/1 S87I, 22/27 (81.5%) S87N, and 16/20 (80.0%) D86N substitutions. For *gyrA*, real-time PCR detected mutations in 64/66 (97.0%) samples classified as non-wild-type by WGS and correctly classified 23/24 (95.8%) concordant WGS wild-type samples.

**Table 2.** Agreement between targeted real-time PCR and WGS for the assessed resistance-associated mutations (n=90)

| Real-time PCR | WGS |  | Performance |  |
|---|---|---|---|---|
|  | Non wild type | Wild type | Positive agreement | Negative agreement |
| ***parC*** |  |  |  |  |
| **SNP** at D86N, S87R/N/I or S88P | 39 | 0 | 81.2%<br>(95% CI 67.4% - 91.1%) | 100%<br>(95% CI 91.6% - 100%) |
| **WT** at D86, S87, S88 | 9 | 42 |  |  |
| ***gyrA*** |  |  |  |  |
| **SNP**: S91F, S91P, S91Y | 64 | 1 | 97.0%<br>(95% CI 89.5% - 99.6%) | 95.8%<br>(95% CI 78.9% - 99.9%) |
| **WT**: S91 | 2 | 23 |  |  |
| **23S – A2059** |  |  |  |  |
| SNP | 0 | 0 | NA | 100%<br>(96.0% - 100%) |
| WT | 0 | 90 |  |  |
| **23S – C2611\*** |  |  |  |  |
| SNP | 1 | 0 | 100%<br>(95% CI 2.5% - 100%) | 100%*<br>(95% CI 95.8% - 100%) |
| WT | 0 | 87 |  |  |

**Abbreviations:** PCR, polymerase chain reaction; WGS, whole genome sequencing; SNP, single nucleotide polymorphism; WT, wild type; CI, confidence interval; NA, not applicable.
\*Excluding the two sample with undetermined result in real-time PCR

### Association between resistance-associated mutations and ciprofloxacin and azithromycin susceptibility

Ciprofloxacin susceptibility results were available for 89/90 isolates with paired real-time PCR and WGS data. Of these, 21 (23.6%) were susceptible, three (3.4%) were intermediate and 65 (73.0%) were resistant (Table 3). Among the 65 ciprofloxacin-resistant isolates, at least one targeted *parC* or *gyrA* mutation was detected in 60 (92.3%) by real-time PCR and in 61 (93.8%) by WGS. Five (7.7%) resistant isolates had no targeted mutation detected by real-time PCR; WGS identified a resistance-associated mutation in one of these.

**Table 3.** Comparison of real-time PCR and whole-genome sequencing results with phenotypic drug susceptibility testing results for ciprofloxacin.

|  | **Ciprofloxacin susceptibility testing\*** |  |  |
|---|---|---|---|
|  | **Susceptible**<br>**(MIC ≤0.032 mg/L)**<br>**(n=21)** | **Intermediate**<br>**(MIC of 0.064 mg/L)**<br>**(n=3)** | **Resistant**<br>**(MIC >0.064 mg/L)**<br>**(n=65)** |
| **Real-time PCR** |  |  |  |
| Wild type at targeted position | 16 (76%) | 1 (33%) | 5 (7.7%) |
| Targeted mutation detected | 5 (24%) | 2 (67%) | 60 (92%) |
| *parC* D86/S87/S88 | 5 (100%) | - | 33 (55%) |
| *gyrA* S91 | 3 (60%) | 2 (100%) | 59 (98%) |
| **WGS** |  |  |  |
| Wild type at targeted position | 16 (76%) | 1 (33%) | 4 (6.2%) |
| Targeted mutation detected | 5 (24%) | 2 (67%) | 61 (94%) |
| *parC* D86/S87/S88 | 5 (100%) | - | 42 (69%) |
| *gyrA* S91/D95 | 2 (40%) | 2 (100%) | 61 (100%) |

\*Minimum inhibitory concentration (MIC) was interpreted according to EUCAST v12.0 (2022)

Four ciprofloxacin-resistant isolates had discordance between phenotype and genotype. Three had ciprofloxacin MICs of 0.5 mg/L and one had an MIC of 32 mg/L (Supplementary table 3). Additional genomic analysis identified a *gyrA* I250M substitution in one of these isolates.

Among the 21 ciprofloxacin-susceptible isolates, *parC* mutations were detected by real-time PCR in five (23.8%), three of which also had a *gyrA* mutation. WGS confirmed the corresponding mutations in all five isolates. Four of these isolates had ciprofloxacin MICs of 0.002 mg/L and one had an MIC of 0.032 mg/L.

For 23S rRNA, real-time PCR and WGS both detected C2611T in one isolate; the remaining evaluable isolates were concordantly wild type at A2059 and C2611. The C2611T-positive isolate had an azithromycin MIC (8 mg/L) above the epidemiological cut-off (>1mg/L). Among the remaining isolates, the median azithromycin MIC was 0.064 mg/L (range, 0.016–1.0 mg/L). One isolate had an MIC of 1.0 mg/L without an A2059 or C2611 mutation detected by either method.

## DISCUSSION

This study characterised the molecular epidemiology and antimicrobial resistance determinants of *N. gonorrhoeae* among symptomatic individuals in Eastern Cape, South Africa. First, substantial NG-MAST sequence-type diversity was observed. Second, the *gyrA* and/or *parC* mutations were relatively more common, as compared to the 23S rRNA mutations. Lastly, real-time PCR performed directly on stored clinical-specimen DNA showed generally good agreement with WGS for the targeted resistance-associated mutations, although genotype and ciprofloxacin phenotype were not fully concordant.

The considerable NG-MAST diversity observed in this study is consistent with previous South African studies reporting heterogeneous gonococcal populations in KwaZulu-Natal, Johannesburg and the private healthcare sector.(22–24) ST 21253 was the most frequently identified sequence type in the present study and was identified in specimens from all five participating facilities and from both men and women. This distribution suggests that ST 21253 was geographically widespread within the study population; however, NG-MAST is based on two highly variable loci and identification of the same sequence type does not establish close genomic relatedness or direct transmission between cases.(8) The findings therefore support the value of combining molecular typing with higher resolution genomic approaches where detailed investigation of transmission or strain relatedness is required.

An important objective of this study was the detection of RAMs directly from *N. gonorrhoeae*-positive DNA extract without culture. Among the 90 specimens with paired WGS results, agreement varied according to the target: positive agreement was 81.2% for *parC* and 97.0% for *gyrA*, while negative agreement was 100% and 95.8%, respectively. Agreement for the 23S rRNA targets was also high, but the C2611T-mutation was only detected in one sample which limits performance interpretation. These findings support molecular testing of clinical specimens as a potentially useful complement to culture-based AMR surveillance, particularly where culture capacity is limited. However, the lower positive agreement observed for *parC* also illustrates that performance is target-dependent and that molecular assays need to be evaluated against locally circulating genetic variants.

Targeted *gyrA* and/or *parC* mutations were detected by real-time PCR in 110/193 samples with evaluable results. The high frequency of fluoroquinolone-associated determinants is consistent with previous reports of substantial ciprofloxacin resistance among gonococcal isolates in South Africa.(22, 24) Among the 65 ciprofloxacin-resistant isolates in the paired analysis, 60 (92.3%) had at least one targeted mutation detected by real-time PCR and 61 (93.8%) by WGS. Nevertheless, four resistant isolates had no targeted resistance-associated mutation detected by either method, while targeted mutations were detected in five phenotypically susceptible isolates. Genotype–phenotype discordance for ciprofloxacin has been described previously, including resistant isolates without commonly recognised resistance-associated mutations in the fluoroquinolone resistance-determining regions.(25) The predictive value of molecular testing therefore depends on the particular genetic determinants included in an assay and their relationship with phenotype, rather than simply on the presence of any mutation in *gyrA* or *parC*.

One of our isolates with an MIC of 0.5 mg/L, also had an additional *gyrA* I250M SNP identified through genomic analysis, although its contribution to ciprofloxacin resistance remains uncertain. These findings highlight the challenges of interpreting phenotypic resistance when recognised genetic determinants are not detected. At the same time, in a global collection of gonococcal isolates, wild-type *gyrA* codon 91 reliably predicted ciprofloxacin susceptibility, highlighting the value of validated molecular markers while emphasising the importance of considering the genetic markers assessed when interpreting genotype–phenotype relationships.(26, 27)

The 23S rRNA macrolide RAMs assessed were uncommon; C2611T was identified by both real-time PCR and WGS in one isolate, which had an azithromycin MIC of 8 mg/L, while no A2059G mutations were detected. The association between this mutation and resistance is influenced by the number of 23S rRNA gene copies carrying the mutation, which could not be established from our results.(12, 28) Furthermore, we did not test for other mechanisms contributing to elevated azithromycin MICs, including alterations affecting the MtrCDE efflux system.(29, 30) Consequently, the low frequency of A2059 and C2611 mutations in this study should not be interpreted as evidence for the absence of other macrolide resistance determinants.

This study has several important limitations. The specimens were obtained from symptomatic individuals attending facilities within a single municipality and may therefore not represent asymptomatic infections or gonococcal populations elsewhere in South Africa. Real-time PCR amplification was unsuccessful in 42 DNA extracts, which may limit the applicability of this approach to specimens with sufficient target DNA. Only 90 of the 193 specimens with resistance-marker real-time PCR results had paired WGS data. The molecular assays assessed predefined mutations and therefore could not identify all genetic mechanisms potentially associated with resistance. In addition, only one C2611T-positive isolate and no A2059-positive isolates were identified, which did not allow for robust assessment of assay performance for these macrolide resistance determinants.

In conclusion, this study demonstrates substantial gonococcal strain diversity and a high frequency of targeted fluoroquinolone resistance-associated mutations among symptomatic individuals in this South African setting. Targeted real-time PCR applied directly to clinical-specimen DNA showed generally good agreement with WGS and may provide a useful complementary approach for molecular AMR surveillance where culture-based surveillance is constrained. However, the observed genotype– phenotype discordance and variation in agreement between molecular targets highlight the importance of continued phenotypic surveillance and local validation of molecular resistance markers. Integrating culture-independent molecular approaches with phenotypic and genomic surveillance could strengthen monitoring of evolving gonococcal antimicrobial resistance.

## Data Availability

All data produced in the present study are available upon reasonable request to the authors

## Statement on Conflict of Interests

All authors declare no competing interests

## Sources of Support

This research received no specific grant from any funding agency in the public, commercial or not-for-profit sectors.

## Acknowledgments

We would like to acknowledge the support provided by the Foundation for Professional Development Research Unit and the Department of Medical Microbiology, University of Pretoria.

## Contributors

MMM conceptualised the study, conducted the laboratory work, analysed the data, and drafted the manuscript. HJ provided laboratory supervision and technical support for the molecular analyses. BK contributed substantially to the bioinformatics and sequence analyses and critically reviewed and revised the manuscript. NM contributed to data analysis and reviewed and revised the manuscript. RP provided overall supervision, contributed to the study design and interpretation of findings, and critically reviewed and revised the manuscript. All authors reviewed and approved the final manuscript.

## Notes

### Competing Interest Statement

The authors have declared no competing interest.

### Author Declarations

The Research Ethics Committee of the Faculty of Health Sciences, University of Pretoria, gave ethical approval for this work (reference number: 42/2024).

