## Supplementary Table 1 for "Molecular typing and characterisation of antimicrobial resistance determinants of *Neisseria gonorrhoeae* among symptomatic individuals in the Eastern Cape, South Africa"

**Supplementary Table 1. SNPs associated with AMR in *Neisseria gonorrhoeae*: by study methods and additional determinants reported by Pathogenwatch**

| **Gene** | **Real-time PCR SNPs** | **WGS SNPs** | *** Additional SNPs reported by Pathogenwatch** |
| --- | --- | --- | --- |
| *parC* | - D86 - S87 - S88 | - D86N - S87N - E91K | - S87R - S87I - S88P |
| *gyrA* | - S91 | - S91F - D95G - D95A | - D95N |
| 23S rRNA | - A2059 - C2611 | - C2611 - A2059 | None |

* **Note:** The additional SNPs column includes literature-reported determinants from Pathogenwatch that were not specifically included in the study's RT-PCR targets or WGS variant list. Inclusion in this table does not imply detection in the study isolates.

**Abbreviations:** SNP, single nucleotide polymorphism; AMR, antimicrobial resistance; WGS, whole genome sequencing
