## Supplementary Table 2 for "Molecular typing and characterisation of antimicrobial resistance determinants of *Neisseria gonorrhoeae* among symptomatic individuals in the Eastern Cape, South Africa"

**Supplementary Table 2. Distribution of *Neisseria gonorrhoeae* sequence types by molecular typing method.**

| Sample ID | Site | Gender | Sample type | ST | Method |
| --- | --- | --- | --- | --- | --- |
| 1008 | Site A | Male | Urine | ST21047 | Real-time PCR |
| 1013 | Site A | Female | Swab | ST26978 | Real-time PCR |
| 1020 | Site A | Male | Urine | ST21200 | WGS |
| 1030 | Site A | Male | Urine | ST21057 | WGS |
| 1033 | Site A | Female | Swab | ST21048 | Real-time PCR |
| 1035 | Site A | Male | Urine | ST21192 | WGS |
| 1036 | Site A | Male | Urine | ST21205 | WGS |
| 1037 | Site A | Male | Urine | ST21261 | Real-time PCR |
| 1038 | Site A | Male | Urine | ST21210 | WGS |
| 1040 | Site A | Female | Swab | ST21257 | Real-time PCR |
| 1052 | Site A | Male | Urine | ST21192 | WGS |
| 1053 | Site A | Female | Swab | ST21271 | Real-time PCR |
| 1060 | Site A | Male | Urine | ST21270 | Real-time PCR |
| 1062 | Site A | Male | Urine | ST19216 | WGS |
| 1065 | Site A | Male | Urine | ST21268 | Real-time PCR |
| 1067 | Site A | Female | Swab | ST21187 | Real-time PCR |
| 1068 | Site A | Male | Urine | ST21187 | WGS |
| 1071 | Site A | Male | Urine | ST21253 | Real-time PCR |
| 1074 | Site A | Female | Swab | ST21315 | WGS |
| 1075 | Site A | Male | Urine | ST21060 | Real-time PCR |
| 1077 | Site A | Male | Urine | ST21047 | WGS |
| 1080 | Site A | Male | Urine | ST21186 | WGS |
| 1087 | Site A | Male | Urine | ST21317 | WGS |
| 1090 | Site A | Male | Urine | ST21186 | WGS |
| 1093 | Site A | Male | Urine | ST21191 | WGS |
| 1099 | Site A | Male | Urine | ST21211 | WGS |
| 1104 | Site A | Male | Urine | ST21193 | Real-time PCR |
| 1105 | Site A | Male | Urine | ST21322 | WGS |
| 1107 | Site A | Male | Urine | ST21190 | WGS |
| 1109 | Site A | Female | Swab | ST21319 | WGS |
| 1116 | Site A | Male | Urine | ST21253 | Real-time PCR |
| 3008 | Site B | Male | Urine | ST1027 | Real-time PCR |
| 3010 | Site B | Male | Urine | ST26979 | Real-time PCR |
| 3015 | Site B | Male | Urine | ST21197 | WGS |
| 3017 | Site B | Male | Urine | ST21201 | WGS |
| 3031 | Site B | Male | Urine | ST21062 | WGS |
| 3044 | Site B | Male | Urine | ST21310 | WGS |
| 3045 | Site B | Male | Urine | ST13743 | Real-time PCR |
| 3051 | Site B | Female | Swab | ST2670 | Real-time PCR |
| 3062 | Site B | Female | Swab | ST21258 | Real-time PCR |
| 3073 | Site B | Male | Urine | ST21062 | Real-time PCR |
| 3075 | Site B | Female | Swab | ST21273 | Real-time PCR |
| 3076 | Site B | Female | Swab | ST21313 | WGS |
| 3082 | Site B | Male | Urine | ST21062 | Real-time PCR |
| 3085 | Site B | Female | Swab | ST21252 | Real-time PCR |
| 3089 | Site B | Female | Swab | ST19216 | Real-time PCR |
| 3090 | Site B | Male | Urine | ST21313 | WGS |
| 3095 | Site B | Male | Urine | ST21260 | Real-time PCR |
| 3096 | Site B | Male | Urine | ST21247 | Real-time PCR |
| 3102 | Site B | Male | Urine | ST21313 | WGS |
| 3104 | Site B | Female | Swab | ST21253 | Real-time PCR |
| 3105 | Site B | Male | Urine | ST21253 | Real-time PCR |
| 3107 | Site B | Male | Urine | ST10107 | WGS |
| 3108 | Site B | Male | Urine | ST21318 | WGS |
| 3115 | Site B | Male | Urine | ST21306 | WGS |
| 3118 | Site B | Male | Urine | ST21254 | Real-time PCR |
| 3121 | Site B | Male | Urine | ST21276 | Real-time PCR |
| 3123 | Site B | Female | Swab | ST21059 | Real-time PCR |
| 3130 | Site B | Male | Urine | ST21254 | WGS |
| 3136 | Site B | Female | Swab | ST689 | WGS |
| 3140 | Site B | Female | Swab | ST21250 | Real-time PCR |
| 3142 | Site B | Male | Urine | ST21200 | Real-time PCR |
| 3144 | Site B | Male | Urine | ST21062 | WGS |
| 3146 | Site B | Male | Urine | ST21055 | WGS |
| 3155 | Site B | Male | Urine | ST21253 | Real-time PCR |
| 3158 | Site B | Male | Urine | ST21275 | Real-time PCR |
| 3159 | Site B | Male | Urine | ST21058 | Real-time PCR |
| 3163 | Site B | Male | Urine | ST21277 | Real-time PCR |
| 3164 | Site B | Male | Urine | ST2670 | Real-time PCR |
| 3165 | Site B | Female | Swab | ST2670 | Real-time PCR |
| 3166 | Site B | Male | Urine | ST21251 | Real-time PCR |
| 3168 | Site B | Male | Urine | ST21277 | Real-time PCR |
| 3171 | Site B | Male | Urine | ST12578 | Real-time PCR |
| 5005 | Site C | Male | Urine | ST21196 | WGS |
| 5006 | Site C | Male | Urine | ST21323 | WGS |
| 5010 | Site C | Male | Urine | ST21323 | WGS |
| 5018 | Site C | Male | Urine | ST21308 | WGS |
| 5025 | Site C | Male | Urine | ST21326 | WGS |
| 5028 | Site C | Female | Swab | ST21207 | WGS |
| 5029 | Site C | Male | Urine | ST21267 | Real-time PCR |
| 5031 | Site C | Male | Urine | ST21211 | Real-time PCR |
| 5033 | Site C | Male | Urine | ST21188 | WGS |
| 5034 | Site C | Female | Swab | ST21320 | WGS |
| 5037 | Site C | Male | Urine | ST21312 | WGS |
| 5039 | Site C | Male | Urine | ST21254 | WGS |
| 5041 | Site C | Male | Urine | ST21062 | WGS |
| 5043 | Site C | Male | Urine | ST21307 | WGS |
| 5044 | Site C | Male | Urine | ST21198 | WGS |
| 5045 | Site C | Male | Urine | ST21312 | WGS |
| 5048 | Site C | Male | Urine | ST21254 | WGS |
| 5050 | Site C | Male | Urine | ST21253 | WGS |
| 5051 | Site C | Male | Urine | ST21054 | WGS |
| 5054 | Site C | Male | Urine | ST21046 | WGS |
| 5057 | Site C | Male | Urine | ST21194 | WGS |
| 5059 | Site C | Male | Urine | ST21266 | Real-time PCR |
| 5060 | Site C | Male | Urine | ST21279 | Real-time PCR |
| 5064 | Site C | Male | Urine | ST21253 | Real-time PCR |
| 5068 | Site C | Male | Urine | ST21263 | Real-time PCR |
| 5070 | Site C | Male | Urine | ST21253 | Real-time PCR |
| 5078 | Site C | Male | Urine | ST21189 | Real-time PCR |
| 5079 | Site C | Female | Swab | ST21189 | Real-time PCR |
| 7001 | Site D | Female | Swab | ST21273 | Real-time PCR |
| 7004 | Site D | Male | Urine | ST2575 | WGS |
| 7005 | Site D | Male | Urine | ST21253 | Real-time PCR |
| 7009 | Site D | Male | Urine | ST21189 | WGS |
| 7011 | Site D | Female | Swab | ST19216 | Real-time PCR |
| 7012 | Site D | Male | Urine | ST26980 | Real-time PCR |
| 7013 | Site D | Male | Urine | ST19216 | WGS |
| 7016 | Site D | Male | Urine | ST21253 | WGS |
| 7019 | Site D | Male | Urine | ST21050 | WGS |
| 7023 | Site D | Male | Urine | ST21051 | WGS |
| 7025 | Site D | Male | Urine | ST2670 | WGS |
| 7031 | Site D | Male | Urine | ST19216 | Real-time PCR |
| 7035 | Site D | Male | Urine | ST21196 | Real-time PCR |
| 7036 | Site D | Male | Urine | ST26981 | Real-time PCR |
| 7039 | Site D | Male | Urine | ST21311 | WGS |
| 7040 | Site D | Male | Urine | ST21255 | WGS |
| 7042 | Site D | Male | Urine | ST21047 | WGS |
| 7046 | Site D | Male | Urine | ST21326 | WGS |
| 7052 | Site D | Male | Urine | ST21321 | WGS |
| 7064 | Site D | Male | Urine | ST21053 | WGS |
| 7067 | Site D | Male | Urine | ST21254 | WGS |
| 7070 | Site D | Male | Urine | ST21314 | WGS |
| 7071 | Site D | Male | Urine | ST2975 | WGS |
| 7072 | Site D | Female | Swab | ST21265 | Real-time PCR |
| 7073 | Site D | Male | Urine | ST21321 | WGS |
| 7077 | Site D | Female | Swab | ST21265 | Real-time PCR |
| 7079 | Site D | Male | Urine | ST21309 | WGS |
| 7083 | Site D | Male | Urine | ST21053 | WGS |
| 7086 | Site D | Male | Urine | ST21253 | WGS |
| 7089 | Site D | Male | Urine | ST21253 | Real-time PCR |
| 7090 | Site D | Female | Swab | ST21253 | Real-time PCR |
| 7092 | Site D | Female | Swab | ST22606 | Real-time PCR |
| 7095 | Site D | Male | Urine | ST21208 | WGS |
| 7096 | Site D | Male | Urine | ST21253 | WGS |
| 7097 | Site D | Male | Urine | ST21253 | Real-time PCR |
| 7101 | Site D | Male | Urine | ST21209 | WGS |
| 7108 | Site D | Male | Urine | ST21195 | WGS |
| 7109 | Site D | Female | Swab | ST21195 | WGS |
| 7110 | Site D | Male | Urine | ST6216 | WGS |
| 7113 | Site D | Male | Urine | ST21324 | WGS |
| 7115 | Site D | Male | Urine | ST21053 | Real-time PCR |
| 7117 | Site D | Female | Swab | ST21253 | Real-time PCR |
| 7124 | Site D | Female | Swab | ST18259 | Real-time PCR |
| 7129 | Site D | Male | Urine | ST21316 | WGS |
| 7135 | Site D | Female | Swab | ST21245 | Real-time PCR |
| 7136 | Site D | Male | Urine | ST21253 | Real-time PCR |
| 7143 | Site D | Male | Urine | ST21199 | WGS |
| 7144 | Site D | Female | Swab | ST21253 | WGS |
| 7146 | Site D | Male | Urine | ST21211 | WGS |
| 7148 | Site D | Male | Urine | ST21045 | WGS |
| 7152 | Site D | Male | Urine | ST21262 | Real-time PCR |
| 7154 | Site D | Female | Swab | ST21274 | Real-time PCR |
| 7156 | Site D | Female | Swab | ST26982 | Real-time PCR |
| 8003 | Site E | Female | Swab | ST26983 | Real-time PCR |
| 8004 | Site E | Male | Urine | ST21279 | Real-time PCR |
| 8005 | Site E | Male | Urine | ST26985 | Real-time PCR |
| 8006 | Site E | Female | Swab | ST26984 | Real-time PCR |
| 8011 | Site E | Female | Swab | ST21272 | Real-time PCR |
| 8014 | Site E | Male | Urine | ST21253 | WGS |
| 8016 | Site E | Male | Urine | ST20367 | WGS |
| 8017 | Site E | Female | Swab | ST20367 | Real-time PCR |
| 8019 | Site E | Male | Urine | ST21195 | Real-time PCR |
| 8020 | Site E | Female | Swab | ST21195 | Real-time PCR |
| 8023 | Site E | Male | Urine | ST21255 | Real-time PCR |
| 8028 | Site E | Male | Urine | ST21253 | WGS |
| 8030 | Site E | Male | Urine | ST21259 | Real-time PCR |
| 8034 | Site E | Male | Urine | ST21202 | WGS |
| 8037 | Site E | Male | Urine | ST21203 | WGS |
| 8039 | Site E | Female | Swab | ST21256 | Real-time PCR |
| 8040 | Site E | Male | Urine | ST21204 | WGS |
| 8041 | Site E | Female | Swab | ST19216 | Real-time PCR |
| 8042 | Site E | Male | Urine | ST2187 | Real-time PCR |
| 8043 | Site E | Female | Swab | ST2187 | Real-time PCR |
| 8046 | Site E | Male | Urine | ST21253 | Real-time PCR |
| 8048 | Site E | Male | Urine | ST21206 | WGS |
| 8050 | Site E | Male | Urine | ST21061 | WGS |
| 8054 | Site E | Female | Swab | ST21052 | WGS |
| 8059 | Site E | Male | Urine | ST21254 | Real-time PCR |
| 8061 | Site E | Male | Urine | ST21253 | Real-time PCR |
| 8062 | Site E | Male | Urine | ST21307 | WGS |
| 8068 | Site E | Female | Swab | ST689 | Real-time PCR |
| 8076 | Site E | Male | Urine | ST21248 | Real-time PCR |
| 8078 | Site E | Male | Urine | ST21278 | Real-time PCR |
| 8081 | Site E | Male | Urine | ST21056 | WGS |
| 8082 | Site E | Male | Urine | ST21253 | WGS |
| 8083 | Site E | Male | Urine | ST689 | Real-time PCR |
| 8084 | Site E | Female | Swab | ST21253 | WGS |
| 8085 | Site E | Male | Urine | ST2187 | WGS |
| 8087 | Site E | Male | Urine | ST21212 | WGS |
| 8088 | Site E | Female | Swab | ST21212 | WGS |
| 8089 | Site E | Male | Urine | ST21325 | WGS |
| 8092 | Site E | Male | Urine | ST21309 | WGS |
| 8096 | Site E | Male | Urine | ST21047 | WGS |
| 8097 | Site E | Male | Urine | ST21253 | WGS |
| 8098 | Site E | Male | Urine | ST21269 | WGS |
| 8099 | Site E | Male | Urine | ST21246 | Real-time PCR |
| 8102 | Site E | Male | Urine | ST21264 | Real-time PCR |
| 8103 | Site E | Male | Urine | ST21253 | Real-time PCR |
| 8110 | Site E | Male | Urine | ST21273 | Real-time PCR |

**Abbreviations:** ST, sequence type; WGS, whole genome sequencing
