## Supplementary Table 3 for "Molecular typing and characterisation of antimicrobial resistance determinants of *Neisseria gonorrhoeae* among symptomatic individuals in the Eastern Cape, South Africa"

**Supplementary Table 3. Discordant fluoroquinolone susceptibility results between AST and molecular methods (RT-PCR and WGS)**

| Isolate | AST: Ciprofloxacin | *MIC (mg/L) | WGS: *parC* mutation | WGS: *gyrA*  mutation | RT-PCR: *parC* mutation | RT-PCR: *gyrA* mutation | ** parC* SNPs | ** gyrA* SNPs |
| --- | --- | --- | --- | --- | --- | --- | --- | --- |
| 1068 | Resistant | 32 | WT | WT | WT | WT | None | None |
| 1074 | Resistant | 0.5 | WT | WT | WT | WT | None | None |
| 5054 | Resistant | 0.5 | WT | WT | WT | WT | None | None |
| 8088 | Resistant | 0.5 | WT | WT | WT | WT | None | *gyrA* I250M |

* AST, antimicrobial susceptibility testing; MIC, minimum inhibitory concentration; WGS, whole-genome sequencing; RT-PCR, real-time polymerase chain reaction; WT, wild type. Ciprofloxacin susceptibility was interpreted according to EUCAST v12.0 breakpoints: susceptible, ≤0.032 mg/L; resistant, >0.064 mg/L. Additional *parC* and *gyrA* SNPs refer to potential variants identified through genomic analysis but not reported by Pathogenwatch.
